# Redefining Hemodynamic Imaging in Stroke: Perfusion Parameter Map Generation from TOF-MRA using Artificial Intelligence

**DOI:** 10.1101/2023.08.22.23294405

**Authors:** Tabea Kossen, Vince I Madai, Felix Lohrke, Orhun Utku Aydin, Jonas Behland, Adam Hilbert, Matthias A Mutke, Martin Bendszus, Jan Sobesky, Dietmar Frey

## Abstract

**Background:** Perfusion assessment in cerebrovascular disease is essential for evaluating cerebral hemodynamics and guides many current treatment decisions. Dynamic susceptibility contrast (DSC) magnetic resonance imaging (MRI) is of great utility to generate perfusion parameter maps, but its reliance on a contrast agent with associated health risks and technical challenges limit its usability. We hypothesized that native Time-of-flight magnetic resonance angiography (TOF-MRA) can be used to generate perfusion parameter maps with an artificial intelligence (AI) method, called generative adversarial network (GAN), offering a contrast-free alternative to DSC-MRI.

**Methods:** We propose an adapted 3D pix2pix GAN that generates common perfusion maps from TOF-MRA images (CBF, CBV, MTT, Tmax). The models are trained on two datasets consisting of 272 patients with acute stroke and steno-occlusive disease. The performance was evaluated by the structural similarity index measure (SSIM), for the acute dataset we calculated the Dice coefficient for lesions with a time-to-maximum (Tmax) >6s.

**Findings:** Our GAN model showed high visual overlap and high performance for all perfusion maps on both the acute stroke dataset (mean SSIM 0.88-092) and data including steno-occlusive disease patients (mean SSIM 0.83–0.98). For lesions of Tmax>6, the median Dice coefficient was 0.49.

**Interpretation:** Our study shows that our AI model can accurately generate perfusion parameter maps from TOF-MRA images, paving the way for clinical utility. We present a non-invasive alternative to contrast agent-based imaging for the assessment of cerebral hemodynamics in patients with cerebrovascular disease. Leveraging TOF-MRA data for the generation of perfusion maps represents a groundbreaking approach in cerebrovascular disease imaging. This method could greatly impact the stratification of patients with cerebrovascular diseases by providing an alternative to contrast agent-based perfusion assessment.

**Funding:** This work has received funding from the European Commission (Horizon2020 grant: PRECISE4Q No. 777107, coordinator: DF) and the German Federal Ministry of Education and Research (Go-Bio grant: PREDICTioN2020 No. 031B0154 lead: DF).

## Introduction

Acute ischemic stroke (AIS) is one of the leading causes of death and disability and requires rapid decision-making to optimize patient outcomes^1^. Traditional patient stratification strategies for recanalization via mechanical intervention often include perfusion imaging. Perfusion imaging, available both for computed tomography (CT) and magnetic resonance imaging (MRI), provides essential information about the severity and extension of ischemia in addition to vessel information^1,2^. In MRI, dynamic susceptibility contrast (DSC) MRI is the most common perfusion-weighted imaging (PWI) technique that requires the application of a gadolinium-based contrast agent^3^.

The use of gadolinium-based contrast agents, however, has raised concerns owing to potential health risks. Gadolinium deposition in organs, also the brain, has been reported, particularly in patients with impaired renal function or repeated exposure to gadolinium^4^. Additionally, PWI faces technical challenges limiting its effectiveness despite decades of dedicated research. One major difficulty is the selection of an appropriate arterial input function (AIF), which can significantly impact the accuracy of deconvolved perfusion parameter maps^5,6^. Additionally, there is considerable variability in the perfusion parameter maps generated by different vendors, even if the same clinically relevant parameter map thresholds are chosen, making it difficult to establish standardized guidelines for interpretation^7^. Consequently, there is an urgent need for alternative techniques for patient stratification without health risks and unreliable post-processing that maintain the required diagnostic information.

Recently, deep learning techniques, particularly generative adversarial networks (GANs), have shown promise in medical imaging applications, especially for image-to-image translations^8,9^. GANs consist of two neural networks, the generator and the discriminator, that compete with each other to enable the generation of high-quality images^10^. In the context of AIS, it has been shown recently that GANs can be employed to generate perfusion parameter maps from perfusion source images without the need for AIF selection and deconvolution, potentially simplifying and improving the diagnostic process^11^.

This work takes this idea a significant step further. We hypothesized that native Time-of-flight (TOF) images contain sufficient hemodynamic information to generate perfusion parameter maps using a GAN. TOF imaging provides a correlate of cerebral hemodynamics via valuable information about vessel configuration, collateral circulation, and the presence of steno-occlusions. Native TOF imaging is free of health risks for patients, as it does not require the administration of any contrast agent.

In this exploratory proof-of-concept study, we thus propose a novel approach that leverages GANs to generate parameter maps directly from native TOF imaging data, bypassing the need for DSC perfusion imaging and associated health risks. By training GANs on TOF imaging data as input and DSC-MRI parameter maps as output, we aimed to generate perfusion maps that can provide valuable insights into the cerebral hemodynamics in AIS. Specifically, we propose a modified 3D pix2pix GAN that translates TOF-MRA images to perfusion parameter maps. Using this approach we generate five commonly used perfusion parameters, i.e. time-to-maximum (Tmax), cerebral blood flow (CBF), cerebral blood volume (CBV), mean transit time (MTT) and time-to-peak (TTP). We train and test our models on two different patient cohorts including acute stroke patients and patients with chronic cerebrovascular disease.

## Materials and Methods

### Data and Pre-processing

In total, 272 patients were included in this study. 200 patients suffered from acute stroke and were enrolled in a study at Heidelberg University Hospital. Imaging was performed with a T2*-weighted gradient-echo EPI sequence with fat supression TR=2220ms, TE=36ms, flip angle 90°, field of view: 240×240mm2, image matrix: 128×128mm, 25-27 slices with a slice thickness of 5 mm and was started simultaneously with bolus injection of a standard dose (0.1mmol/kg) of an intravenous gadolinium-based contrast agent on 3 Tesla MRI systems (Magnetom Verio, TIM Trio and Magnetom Prisma; Siemens Healthcare, Erlangen, Germany). 50 to 75 dynamic measurements were performed (including at least eight prebolus measurements). Bolus and prebolus were injected with a pneumatically driven injection pump at an injection rate of 5ml/s. The study protocol for this retrospective analysis of our prospectively established stroke database was approved by the ethics committee of Heidelberg University and patient informed consent was waived.

For the data from Heidelberg, DSC data were post-processed with Olea Sphere® (Olea Medical®, La Ciotat, France), automatic motion correction was applied. Raw DSC images were used to calculate perfusion maps of TTP from the tissue response curve. Maps of CBF, CBV, MTT, and Tmax were created by deconvolution of a regional concentration time curve with an AIF. Block-circulant singular value decomposition deconvolution was applied. The AIF was detected automatically and then visually inspected by a neuroradiology expert (MAM, >6 years experience in perfusion imaging) and only in two cases the automatically detected AIF needed to be manually corrected.

72 patients with steno-occlusive disease were included from the PEGASUS study^12^. 80 whole-brain images were recorded using a single-shot FID-EPI sequence (TR=1390ms, TE=29ms, voxel size: 1.8×1.8×5mm^3^) after injection of 5ml Gadovist® (Gadobutrol, 1M, Bayer Schering Pharma AG, Berlin, Germany) followed by 25ml saline flush by a power injector (Spectris, Medrad Inc., Warrendale PA, USA) at a rate of 5ml/s. The acquisition time was 1:54 minutes. All patients gave their written informed consent and the study has been authorized by the ethical review committee of Charité - Universitätsmedizin Berlin. DSC post-processing was performed blinded to clinical outcome.

For PEGASUS patients, DSC data were post-processed with the PGui software (Version 1.0, provided for research purposes by the Center for functional neuroimaging, Aarhus University, Denmark). Motion correction was not available. Raw DSC images were used to calculate perfusion maps of TTP from the tissue response curve. Maps of CBF, CBV, MTT, and Tmax were created by deconvolution of a regional concentration time curve with an AIF. Parametric deconvolution was applied^13^. For each patient, an AIF was determined by a junior rater (JB, two years of experience in perfusion imaging) by manual selection of three or four intravascular voxels of the MCA M2 segment contralateral to the side of stenosis minimizing partial volume effects and bolus delay. The AIF shape was visually assessed for peak sharpness, bolus peak time and amplitude width^5,14^. The AIFs were inspected by a senior rater (VIM, >12 years of experience in perfusion imaging).

The TOF-MRA scans were conducted on a clinical 3T whole-body system (Magnetom Trio, Siemens Healthcare, Erlangen, Germany) utilizing a 12-channel receive radiofrequency coil (Siemens Healthcare) for head imaging. For both studies the parameters were: voxel size=(0.5x0.5x0.7)mm^3^; matrix size: 312x384x127; TR/TE = 22ms/3.86ms; acquisition time: 3:50min, flip angle=18°.

For both datasets skullstripping was applied using SynthStrip^15^. Non-uniformity correction was performed on the TOF-MRA images and the perfusion parameter maps were co-registered using the VINCI software package (https://vinci.sf.mpg.de/). All images were resized to a dimension of 256x256x128 and normalized between -1 and +1. Both data sets were split into training (acute stroke data: 140, PEGASUS: 51), validation (acute stroke data: 40, PEGASUS: 13) and test (acute stroke data: 20, PEGASUS: 8) set respectively. The models were trained on the training set with hyperparameters that were determined based on the validation set. The overall generalizable performance was estimated by the performance on the test set.

### Network architecture

The proposed network architecture is a 3D adaptation of the pix2pix GAN^16^. Like the original pix2pix GAN our GAN consisted of two networks: the generator *G* and the discriminator *D*. The generator’s task was to synthesize a 3D perfusion parameter map based on the corresponding TOF-MRA. In contrast to that, the discriminator tried to distinguish between the generated data volume and the ground truth, along with the real TOF-MRA volume (Figure 1). The implementation details of the GAN can be found in appendix 1.

**Figure 1:**
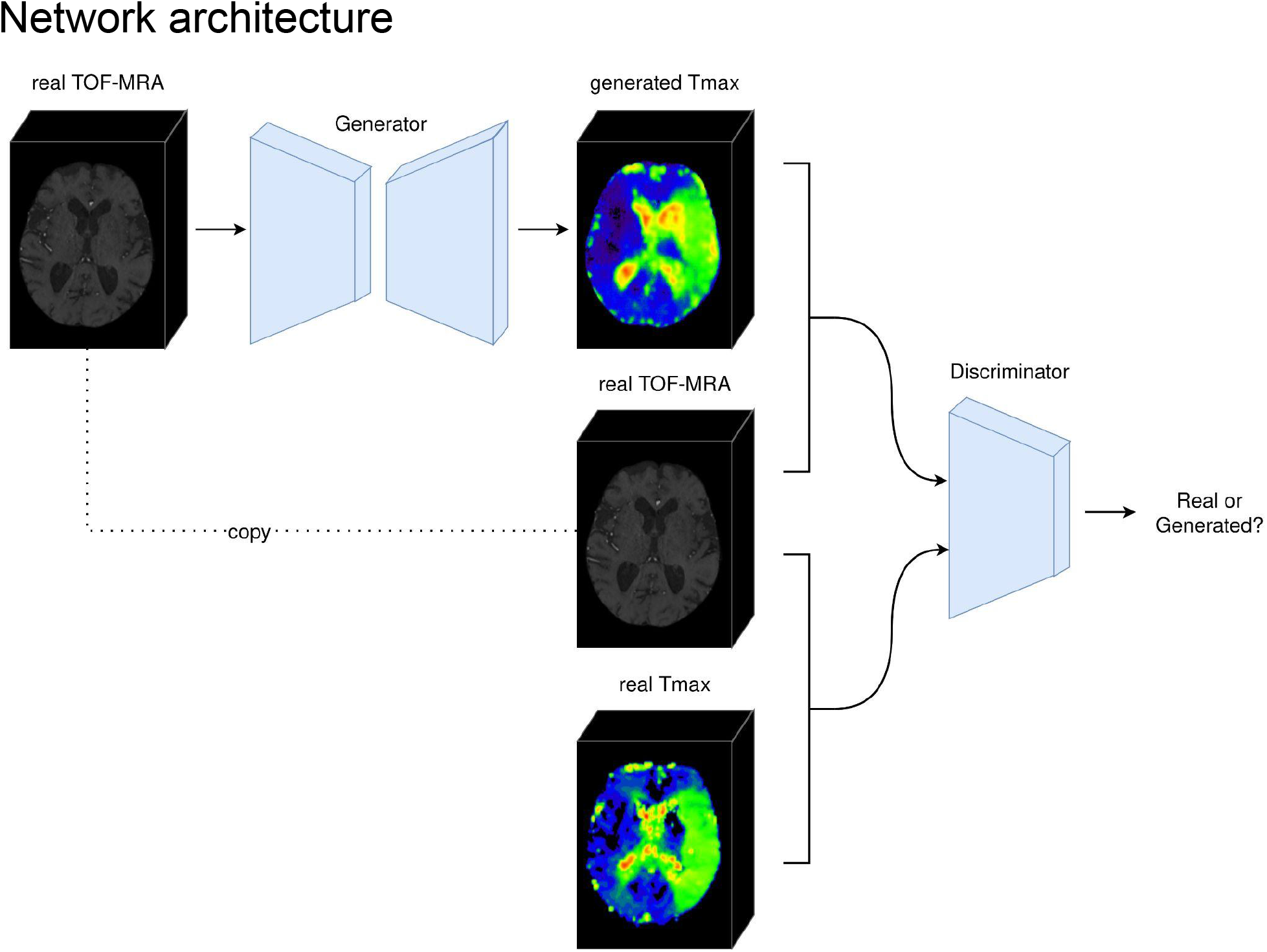
Proposed 3D GAN architecture. The generator takes in 3D TOF-MRA volumes as an input and creates perfusion parameter maps such as Tmax from it. The discriminator’s task is to differentiate between the true TOF-MRA and the true Tmax or the generated Tmax.

### Training

For each dataset, five GANs were trained corresponding to the five perfusion parameter maps (Tmax, CBF, CBV, MTT, TTP). Since the PEGASUS dataset was smaller than the acute dataset, the network parameters of the respective perfusion map from the acute data were used for pre-initialization of the network trained on PEGASUS data.

All models were trained for 200 epochs at which all models had reached convergence. Due to computational restrictions, the batch size was set to 1. The following hyperparameters were varied and selected according to the performance on the validation set. The hyperparameters can be found in appendix 2. The code for the 3D GAN was implemented in PyTorch and is openly available^17^. All models were trained on a TESLA V100 GPU (NVIDIA Corporation, Santa Clara, CA, USA).

### Performance evaluation

After visual inspection, four metrics were calculated to measure the similarity of the generated perfusion parameter maps and the parameter maps derived from the DSC image. The first metric is the structural similarity index measure (SSIM) which combines differences in luminance, contrast and structure. The second metric peak signal-to-noise ratio (PSNR) computes the ratio between the maximal possible signal power and the noise power that is entailed in the data. In both metrics, higher values correspond to increased similarity between the images. The last two metrics mean absolute error (MAE) or L1 norm and normalized root mean squared error (NRMSE) are error metrics. Thus, more similar images lead to lower metrics. Additional details about the metrics are included in appendix 3.

To quantify a clinically relevant alignment between the real and generated perfusion maps, an additional segmentation based analysis was performed measuring the overlap of the hypoperfused areas. A junior rater (OUA) with 4 years of experience in stroke imaging evaluated pairs of perfusion maps (Tmax) for 20 acute stroke patients in a pseudoanonymized and randomized manner, amounting to 40 maps in total (20 real and 20 generated). The process of segmentation consisted of two steps: thresholding above a Tmax value of 6 as a preliminary segmentation step, followed by the manual removal of artifacts found outside of the hypoperfused area. The Dice coefficient was calculated for quantitative assessment of overlap between original and generated Tmax images.

## Results

Visual inspection showed great similarities between the generated and the DSC-derived perfusion parameter maps for acute stroke patients (see Figure 2 and 3). Particularly, in Figure 2, the hypoperfused areas, visualized by generated Tmax, have great overlap with the ground truth. The quantitative analysis using image-based similarity metrics confirmed this showing a high performance for all perfusion parameter maps with SSIM>0.87, PSNR>28, MAE<0.04 and NRMSE<0.4 (see Table 1).

**Figure 2:**
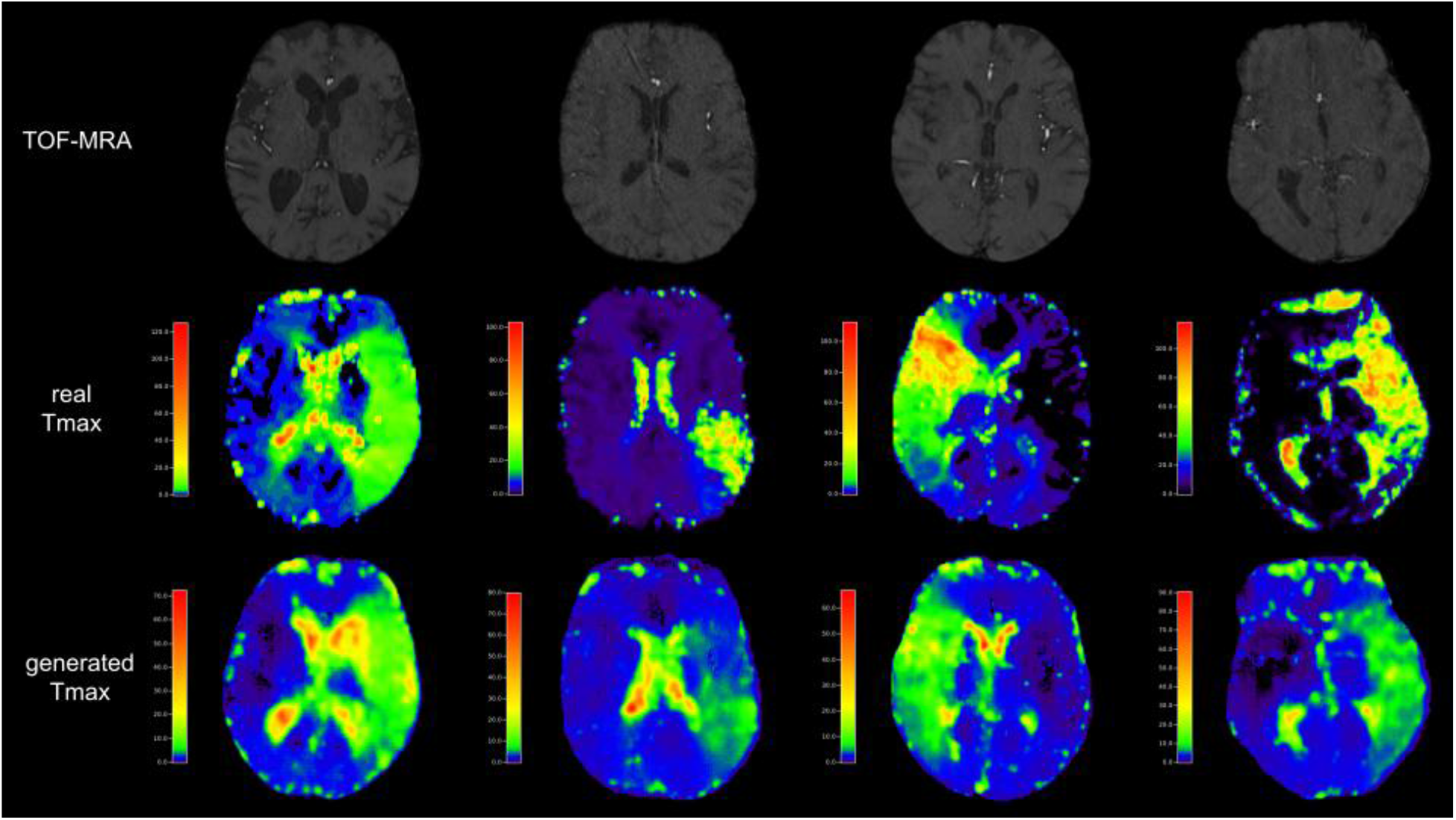
Generated Tmax (bottom row) from TOF-MRA images (top row) compared to the DSC-derived Tmax (middle row) for the acute stroke dataset. The example in the first column shows an image with the highest performance, the ones in the second and third column average performance and the last one the overall lowest performance. For all categories, however, our model was able to capture the relevant hypoperfused area.

**Figure 3:**
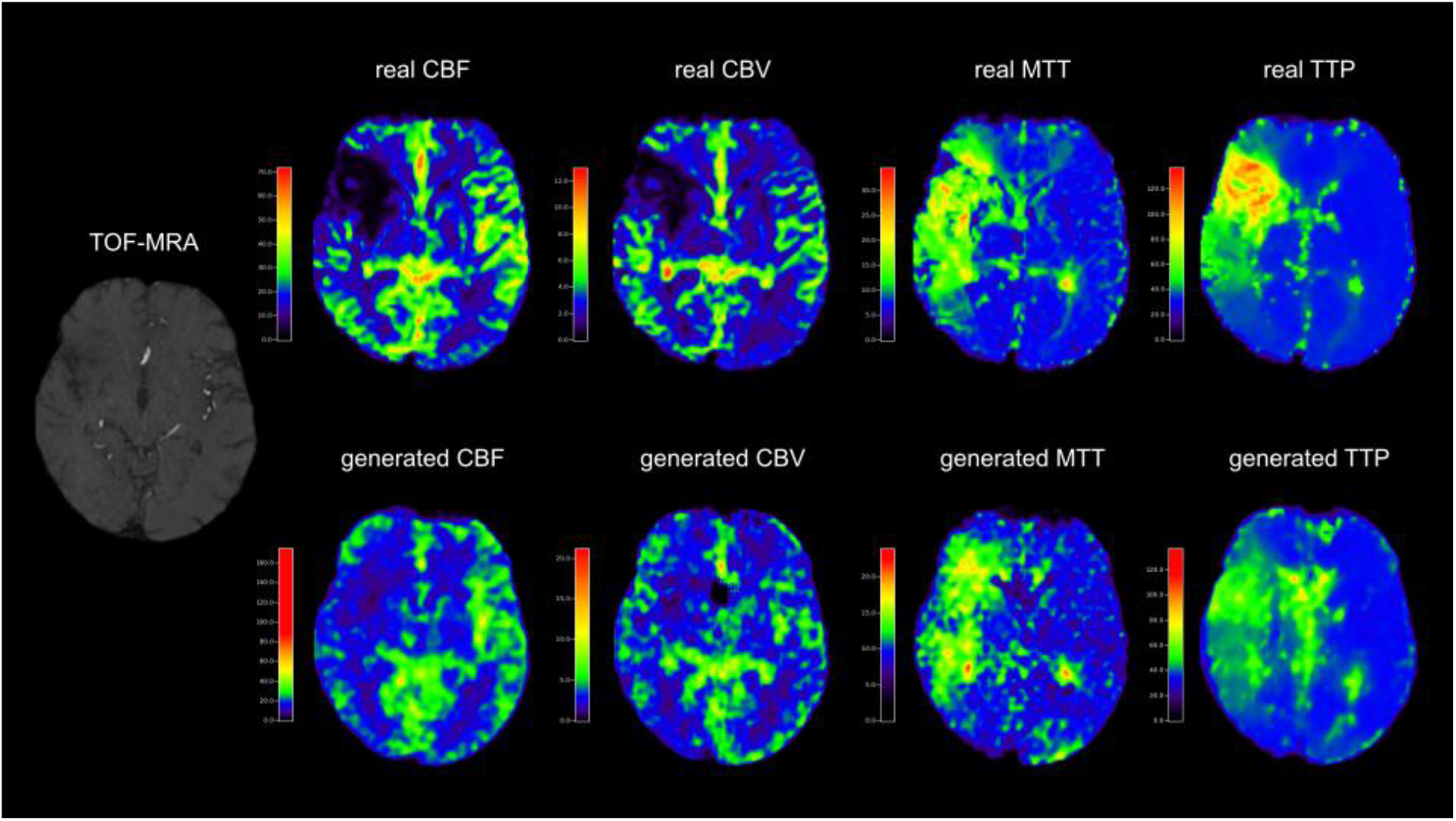
Generated parameter maps CBF, CBV, MTT and TTP (bottom row) from TOF-MRA images (left) compared to the DSC-derived maps (top row) for the acute stroke dataset. The generated perfusion maps show high similarity to the ground truth.

**Table 1:** Mean performance metrics for evaluating the similarity between the ground truth and the synthesized parameter maps for the acute stroke and PEGASUS dataset. The standard deviations across patients are shown in brackets.

|  | SSIM | PSNR | MAE | NRMSE |
| --- | --- | --- | --- | --- |
| Acute stroke data: |  |  |  |  |
| Tmax | 0.918 (0.033) | 29.618 (3.220) | 0.022 (0.010) | 0.202 (0.089) |
| CBF | 0.876 (0.030) | 28.480 (2.595) | 0.036 (0.014) | 0.272 (0.126) |
| CBV | 0.905 (0.016) | 30.889 (2.016) | 0.025 (0.007) | 0.141 (0.039) |
| MTT | 0.905 (0.031) | 30.477 (2.541) | 0.029 (0.010) | 0.191 (0.085) |
| TTP | 0.903 (0.040) | 28.527 (3.324) | 0.036 (0.017) | 0.374 (0.213) |
| PEGASUS: |  |  |  |  |
| Tmax | 0.973 (0.014) | 37.417 (2.370) | 0.008 (0.003) | 0.028 (0.009) |
| CBF | 0.977 (0.017) | 38.112 (5.508) | 0.009 (0.005) | 0.030 (0.019) |
| CBV | 0.948 (0.021) | 34.208 (3.270) | 0.017 (0.008) | 0.043 (0.017) |
| MTT | 0.832 (0.411) | 23.615 (3.142) | 0.050 (0.021) | 0.149 (0.061) |
| TTP | 0.882 (0.039) | 27.202 (3.313) | 0.034 (0.013) | 0.099 (0.040) |

The additional segmentation-based analysis demonstrated a moderate degree of overlap between the real and generated perfusion maps (see Figure 4). One patient was excluded from this analysis due to the absence of a discernible lesion in the real perfusion map. For the remaining 19 patients, we report a median Dice value of 0.490 and a mean Dice value of 0.477, with the interquartile range spanning from 0.386 (25th percentile) to 0.656 (75th percentile). Notably, in two patients, the generated hypoperfused area appeared on the contralateral side relative to the real hypoperfused area resulting in no overlap between real and generated lesions.

**Figure 4:**
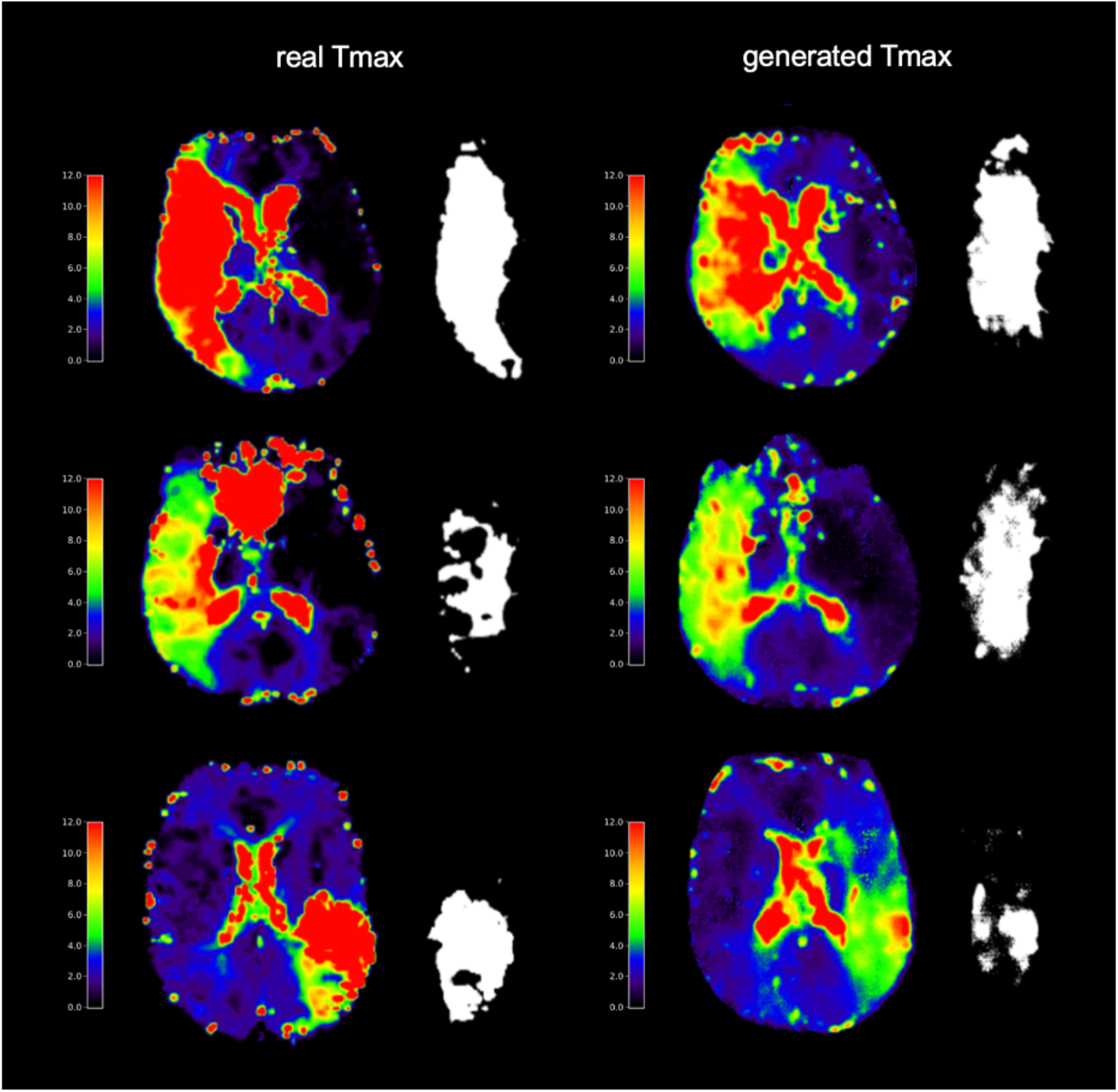
Segmentation-based overlap analysis of real and generated Tmax perfusion maps. From top to bottom the Dice coefficients comparing the manual segmentations (in white) are 0.76, 0.40 and 0.37. The analysis depicts the discrepancy between the visually assessed similarity and the quantified overlap of real and generated lesions with a Tmax>6.

For the PEGASUS dataset including patients with chronic steno-occlusive cerebrovascular disease, the generated perfusion maps also show high similarities to the ground truth visually (see Appendix 4). This was reflected in the metrics, showing performances for all parameter maps with SSIM (structural similarity index measure) >0.83, PSNR (peak signal-to-noise ratio) >23, MAE (mean absolute error) <=0.5 and NRMSE (normalized root mean squared error) <0.2 (see Table 1).

## Discussion

We present a GAN-based AI modeling approach for generating DSC-type perfusion parameter maps (such as Tmax, CBF, CBV, MTT, and TTP) directly from native, non contrast-enhanced TOF-MRA images. Our method aimed to provide a non-invasive alternative to DSC imaging for the assessment of cerebral hemodynamics in patients with acute stroke and steno-occlusive disease. By utilizing a 3D adaptation of a pix2pix GAN deep learning architecture, our model effectively synthesized realistic perfusion parameter maps, demonstrating a strong correlation with the ground truth parameter maps derived from DSC images. The results of this study strongly indicate that TOF-MRA-derived perfusion maps can potentially serve as a meaningful alternative to DSC imaging in certain clinical scenarios to guide patient stratification in cerebrovascular disease. A TOF-MRA based approach would save scanning time and scanning costs, would avoid contrast-agent related health risks, and would also be applicable in cases where contrast administration or longer acquisition times may be contraindicated. Specifically in chronic steno-occlusive disease, this method has the potential of becoming a screening tool allowing repeated assessments of hemodynamics over time.

Our study included two datasets, one from Heidelberg University Hospital comprising acute stroke patients and another from the PEGASUS study, which included patients with steno-occlusive disease. The performance metrics - SSIM, PSNR, MAE, and NRMSE - demonstrated a high degree of similarity between the generated and ground truth perfusion parameter maps for both datasets. Visually, there is also a striking similarity of the main areas affected by hemodynamic changes such as hypoperfusion.

In chronic steno-occlusive disease, our method has the potential of becoming a screening tool allowing repeated assessments of hemodynamics over time. This could address some of the limitations currently faced in the field of cerebrovascular disease monitoring. Traditional imaging-based screening techniques focus only on the morphological aspects of the vessels, which may not always provide a comprehensive assessment of hemodynamics. Furthermore, these imaging techniques are associated with ionizing radiation exposure, the need for contrast agents that may cause allergic reactions or nephrotoxicity, or have relatively high costs^18^. The noninvasive and dynamic nature of our proposed method offers a promising alternative to these current techniques, as it can provide real-time information without the aforementioned disadvantages. This approach could facilitate serial monitoring of patients with dynamic chronic steno-occlusive disease, enabling clinicians to better assess disease progression. Additionally, the ability to monitor hemodynamics in a longitudinal manner could aid in identifying patients at increased risk for future cerebrovascular events, allowing for early intervention and potentially improving overall patient outcomes. However, further research is needed to validate this method and explore its full potential in the context of chronic cerebrovascular disease monitoring.

In acute stroke, the overall quantitative similarity metrics were also highly promising. Yet, the moderate quantitative overlap in the segmentation-based analysis and two cases of flipped lesion locations indicate that providing clinically relevant hypoperfused areas with a Tmax value above 6 clearly needs further research and refinement. Here, we would like to emphasize that our study serves as an exploratory analysis, a proof-of-concept. Further validation of our approach, particularly in the context of patient stratification in acute stroke, is crucial, both from a clinical as well as from a technical point of view.

Clinically, major acute stroke studies, such as the DEFUSE 3, DAWN and EXTEND IA studies, have employed DSC-MRI or CT perfusion to assess perfusion parameters and identify patients who may benefit from reperfusion therapies^18–20^. Current guidelines allow stratification of patients based on advanced perfusion imaging under certain conditions^21,22^. Our noninvasive approach may offer a unique alternative by using vessel status to produce perfusion maps. While our methodology is currently dependent on TOF-MRA, the general concept of utilizing vessel information to generate perfusion maps can be adapted to the CTA domain and potentially contribute to the development of patient stratification strategies that are more broadly and easily applicable. Since vessel imaging in the form of either CTA or TOF-MRA is routinely available in acute stroke setting there is an opportunity to reassess existing retrospective clinical trial data and to analyze the capability of our methodology to guide patient stratification in acute stroke. Naturally, the performance of our method needs to be rigorously evaluated in prospective clinical trials to establish its reliability and clinical impact.

From a technical point of view, we envision several paths how the clinically relevant Tmax lesion generation performance can be enhanced in future studies. Next to testing different GAN architectures, there is also an exciting avenue for future research incorporating additional clinically relevant information in a multimodal modeling approach. The GAN model could potentially yield more accurate and clinically relevant results by integrating clinical features, such as patient demographics, stroke severity, and vascular risk factors. To further increase the overlap of the generated and real maps, segmentation masks of hypoperfused areas based on clinically relevant perfusion threshold values can be provided as additional input to the model to focus the training on the hypoperfused areas. Additionally, other imaging modalities like diffusion-weighted imaging (DWI) and fluid-attenuated inversion recovery (FLAIR) could be integrated to enhance the model’s ability to account for individual patient characteristics. This is especially interesting, since TOF-imaging is often part of stroke protocols or can easily be added. For example, there is the possibility to integrate the DWI/FLAIR-mismatch paradigm for patient stratification into the model. Taken together, the integration of multimodal data could also aid in the development of more personalized and precise patient stratification strategies in AIS, ultimately improving patient outcomes in the context of acute stroke management.

Our work is the first work deriving perfusion parameters from non-contrast-enhanced TOF-MRA images. Previous studies have demonstrated that deep learning methods can be utilized to create perfusion parameter maps based on DSC-MRI for AIS patients as a complementary approach to AIF selection and deconvolution^11,23^. Similarly, Asaduddin et al.^24^ generated perfusion-relevant parameters as well as the vessel architecture from a single contrast-enhanced scan, i.e., dynamic angiography MRI. Although these approaches showcase the success of deep learning-based image-to-image translation, they rely on contrast-enhancement. In contrast to this, we utilized native TOF-MRA to reduce scanning time and health risks. Our work shows that AI digital health technology has the potential to leapfrog stratification of acute stroke patients.

There are several limitations to this study. First, the sample size of the PEGASUS dataset was relatively small compared to the Heidelberg dataset, which may have affected the model’s performance in generalizing to a broader patient population. Second, the study utilized retrospective and mono-centric data, which could introduce potential biases. Third, the results of this study are exploratory in nature and need further experimentation and validation for transferability to clinical workflows. Fourth, the vascular imaging was performed using only TOF-MRA. Adaptation of our methodology to CTA-based generation of corresponding CT perfusion maps is likely feasible but needs further exploration. Lastly, due to computational constraints we did not translate more recent deep learning architectures to 3D such as DP-GAN or image-to-image latent diffusion models^25,26^. Future studies should consider using larger cohorts and, ultimately, prospective evaluations to validate the findings and further explore the clinical utility of TOF-MRA-derived perfusion maps.

## Conclusion

In conclusion, our exploratory study demonstrates that a 3D GAN model can accurately generate perfusion parameter maps from native TOF-MRA images. Our results highlight the importance of vascular status for patient stratification and may pave the way for a non-invasive alternative to contrast agent-based imaging for the assessment of cerebral hemodynamics in patients with acute stroke and steno-occlusive disease. This could potentially revolutionize the clinical management of patients affected by these conditions. Our study urgently warrants research to improve and validate these findings to explore the potential clinical application of our method.

## Supporting information

Supplementary_Material_GAN_TOF_perfusion_manuscript

## Data Availability

The datasets presented in this article are not readily available because data protection laws prohibit sharing of the PEGASUS and acute stroke datasets at the current time point. Requests to access these datasets should be directed to the Ethical Review Committee of Charite Universitatsmedizin Berlin.

## Acknowledgements

Computation has been performed on the HPC for Research cluster of the Berlin Institute of Health.

## Footnotes

1 WHO EMRO Stroke, Cerebrovascular Accident | Health Topics. Available online at: http://www.emro.who.int/health-topics/stroke-cerebrovascular-accident/index.html.

