## Supplementary_Material_GAN_TOF_perfusion_manuscript for "Redefining Hemodynamic Imaging in Stroke: Perfusion Parameter Map Generation from TOF-MRA using Artificial Intelligence"

### Appendix 1: Implementation details of GAN

As in the original pix2pix GAN the overall objective function is:

$$L_{cGAN}(G,D)=E_{x,y}\left[\log(D(x,y))\right]+E_{x,z}\left[\log(1-D(x,G(x,z)))\right]$$

where $x$ is the input image (TOF-MRA) and $y$ the output image (e.g. Tmax) and $z$ a noise vector. The generator tried to maximize this objective by generating a realistic looking image to which the discriminator assigned a high probability for being real. In comparison to that, the discriminator tried to minimize the objective function by assigning high probabilities to the real images and low ones to the synthesized images. The pix2pix GAN does not directly take in the noise $z$ but introduces randomness in the network by using dropout in the generator.

The generator's loss function was split into two parts. The first loss was the adversarial loss which incorporated the discriminator's feedback on the synthesized images. The second part penalized the deviation to the original images directly by calculating for example the L1 loss between the real and generated image:

$$loss_{L1}=\|y-G(x,z)\|_1$$

For the generator's architecture, we utilized a 3D U-Net[27] that took in the entire 3D TOF-MRA. The TOF-MRA volumes were fed through 7 down- and upsampling layers. Each downsampling layer consisted of a convolutional layer, instance normalization as well as a LeakyReLU activation function with slope 0.2. The upsampling layers contained ConvTranspose layers followed by instance normalization, a dropout layer and a ReLU activation. After the last convolutional block, a tanh was applied to the network's output.

The discriminator's architecture was a modified 3D version of the suggested PatchGAN discriminator by Isola et al.[16]. It contained four convolutional layers with instance normalization and a leaky ReLU with slope 0.2. At the end of the network another convolutional layer with a sigmoid activation function was added. Both generator and discriminator used the kernel size of 4 with strides of 2.

### Appendix 2: Selected hyperparameters

Both the generator and discriminator used the Adam optimizer with $\beta_1$ = 0.5 and $\beta_2$ = 0.999 and a learning rate of 0.0001. Dropout was 0. The GAN generating Tmax used the L1 norm for the reconstruction loss and normal weight initialization. The model for TTP also applied the L1 norm but used Xavier normalization as initial weights. The other models used a combination of L1 norm and the structural similarity index measure (SSIM) and normal weight initialization.

### Appendix 3: Performance metrics specification

The structural similarity index measure (SSIM) is defined as:

$$\mathrm{SSIM}(y,\hat{y}) = \frac{(2\mu_y\mu_{\hat{y}} + c_1)(2\sigma_{y\hat{y}} + c_2)}{(\mu_y^2 + \mu_{\hat{y}}^2 + c_1)(\sigma_y^2 + \sigma_{\hat{y}}^2 + c_2)},$$

where $\mu_y$ and $\mu_{\hat{y}}$ are the average values of y and ŷ respectively, $\sigma_y$ the variance and $\sigma_{y\hat{y}}$ the covariance. $c_1$ and $c_2$ are constants for stabilization and defined as $c_1=(k_1L)^2$ and $c_2=(k_2L)^2$ with L being the dynamic range of the pixel values and $k_1,k_2 \ll 1$ small constants. The higher the SSIM, the more similar are the images to each other with 1 denoting the highest similarity.

The second metric peak signal-to-noise ratio (PSNR) computes the ratio between the maximal possible signal power and the noise power that is entailed in the data:

$$\mathrm{PSNR} = 10\log\left(\frac{\mathrm{MAX}_I}{\mathrm{MSE}}\right)$$

with $\mathrm{MAX}_I$ being the maximal possible pixel/voxel value.

The MAE measures the average absolute of the error between the real image y and the generated image ŷ and is defined voxel-wise:

$$\mathrm{MAE} = \frac{1}{n}\sum_{i=1}^{n}|y_i - \hat{y}_i|$$

The NRMSE is calculated by the root mean squared error normalized by average euclidean norm of the real image y:

$$\mathrm{NRMSE} = \frac{\mathrm{RMSE}}{\sqrt{\frac{1}{n}\sum_{i=1}^{n} y_i^2}}$$

with

$$\mathrm{RMSE} = \sqrt{\frac{1}{n}\sum_{i=1}^{n}(y_i - \hat{y}_i)^2}$$

#### Appendix 4: Visual results on PEGASUS dataset

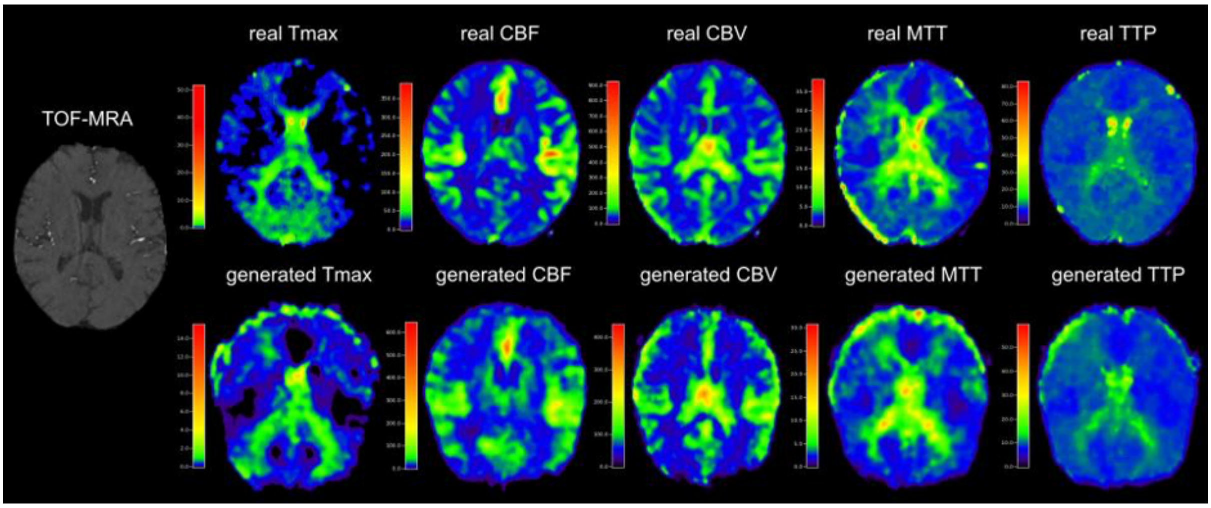


**Figure 5**: Generated parameter maps (bottom row) from TOF-MRA images (left) compared to the DSC-derived maps (top row) for the PEGASUS dataset. The generated perfusion maps show high similarity to the ground truth.
